# Software Application Profile: SUMnlmr, an R package that facilitates flexible and reproducible non-linear Mendelian randomisation analyses

**DOI:** 10.1101/2021.12.10.21267623

**Authors:** Amy M. Mason, Stephen Burgess

**Affiliations:** British Heart Foundation Cardiovascular Epidemiology Unit, Department of Public Health and Primary Care, University of Cambridge, Cambridge, UK; MRC Biostatistics Unit, University of Cambridge, Cambridge, UK

**Keywords:** causal inference, fractional polynomials, piecewise linear, walled garden

## Abstract

**Motivation:** Mendelian randomisation methods that estimate non-linear exposure—outcome relationships typically require individual-level data. This package implements non-linear Mendelian randomisation methods using stratified summarised data, facilitating analyses where individual-level data cannot easily be shared, and additionally increasing reproducibility as summarised data can be reported. Dependence on summarised data means the methods are independent of the form of the individual-level data, increasing flexibility to different outcome types (such as continuous, binary, or time-to-event outcomes).

**Implementation:** SUMnlmr is available as an R package (version 3.1.0 or higher).

**General features:** The package implements the previously proposed fractional polynomial and piecewise linear methods on stratified summarised data that can either be estimated from individual-level data using the package or supplied by a collaborator. It constructs plots to visualise the estimated exposure— outcome relationship, and provides statistics to assess preference for a non-linear model over a linear model.

**Availability:** The package is freely available from GitHub [https://github.com/amymariemason/SUMnlmr].

## Introduction

Mendelian randomisation is a technique for investigating causal relationships using genetic variants as instrumental variables (1). The feasibility and popularity of the approach has increased substantially with the availability of summarised data from genome-wide association studies (GWAS) (2). Methods for the analysis of summarised data typically require the assumption of a linear causal relationship between the exposure and outcome. However, the true relationship may be non-linear – as is the case for body mass index and all-cause mortality (3).

Several methods have been proposed for estimating non-linear causal relationships using instrumental variables, including semiparametric methods (4) and control function methods (5). These methods require access to the full individual-level dataset of genetic variants, outcome, and exposure to fit a non-linear model. This is a major practical barrier to their implementation. The sharing of individual-level data is fraught with security and privacy concerns. Availability of summarised data has democratised genetic epidemiology by enabling researchers from around the world to perform analyses using world-leading datasets. This has catalysed the formation of large global consortia, such as the Covid-19 Host Genetics Initiative (6), enabling authoritative downstream analyses with large sample sizes. The use of publicly-available summarised data also enhances reproducibility, as analyses can easily be replicated.

We here introduce a statistical software package, **SUMnlmr**, that implements the semiparametric methods of Staley and Burgess (4) in a flexible way by splitting the analysis into two stages which can be undertaken independently by separate analysts. First, genetic associations with the exposure and outcome are estimated within strata of the sample. This stage requires access to individual-level data. Secondly, the resulting stratified summarised data are used to fit an appropriate non-linear statistical model. This allows a researcher with access to the stratified summarised data to perform non-linear Mendelian randomisation methods without requiring the individual-level data. A further advantage of this approach is that by standardising inputs, the same version of the non-linear method can be used whatever the original form of the data, as the stratified summarised data have the same form whether the outcome is binary, continuous, or time-to-event: any analytic choices specific to the data (such as the use of logistic or Cox regression) are incorporated into the calculation of the stratified summarised data.

The stratified summarised data required for this approach differ from the summarised data typically reported by GWAS, limiting the application of the method. However, compared to other methods requiring full access to individual-level data, we believe that this approach has substantial practical advantages that make these analyses more accessible, transparent, and reproducible.

In this paper, we first introduce the methods and the software package to implement them. We then perform a simulation study showing the summarised data versions of the methods provide essentially identical estimates to the individual-level data versions for any given choice of model. Finally, we apply the methods to investigate the shape of the causal relationship between low-density lipoprotein (LDL) cholesterol and coronary artery disease risk using data from UK Biobank.

### Implementation

#### Methods

We here provide a brief overview of the methods; a detailed description is available elsewhere (4).

The first step of the approach is to stratify the sample. We cannot stratify on the exposure directly, as it is on the causal pathway from the genetic variants to the outcome, and hence stratification would induce collider bias (7). Instead, we stratify on the ‘residual exposure’, defined as the residual from regression of the exposure on the genetic variants (8). The exposure is influenced by the genetic variants, and so the distribution of the genetic variants would not be uniform within strata based on the exposure – instead, genetic variants associated with lower values of the exposure would be more common in strata with low values of the exposure. In contrast, the residual exposure is independent of the genetic variants, and so the distribution of the genetic variants should be uniform across strata based on the residual exposure.

For simplicity, we assume that there is a single genetic instrument. If there are multiple genetic variants, a single instrument can be obtained by constructing a genetic score (9). Within each stratum, we calculate associations of the genetic instrument with the exposure and with the outcome. The beta-coefficients and standard errors representing these associations are our stratified summarised data. To fit the non-linear models, we also calculate the mean value of the exposure in each stratum, and the 10th and 90th percentiles of the exposure. In the lowest and highest strata, to avoid excessive extrapolation, we instead calculate the 20th and 80th percentiles. These stratified summarised data (see Table 1) can be obtained from individual-level data using the *create_summary_data* function in the **SUMnlmr** package.

**Table 1.** Example of stratified summarised data. Within each stratum, we calculate the genetic association with the exposure [beta-coefficient and standard error (SE)], the genetic association with the outcome (beta-coefficient and SE), and the mean, 10th percentile (lower), and 90th percentile (upper) of the exposure in that stratum. To avoid excessive extrapolation, in the lowest and highest strata we instead calculate the 20th and 80th percentiles.

| Stratum index | Genetic association<br>with exposure |  | Genetic association<br>with outcome |  | Exposure values |  |  |
| --- | --- | --- | --- | --- | --- | --- | --- |
|  | Beta | SE | Beta | SE | Mean | Lower | Upper |
| 1 | 0.253 | 0.006 | 0.154 | 0.049 | 2.45 | 2.30 | 2.70 |
| 2 | 0.227 | 0.004 | 0.249 | 0.050 | 2.75 | 2.53 | 2.93 |
| 3 | 0.226 | 0.003 | 0.205 | 0.048 | 2.97 | 2.74 | 3.11 |
| 4 | 0.234 | 0.002 | 0.197 | 0.048 | 3.14 | 2.92 | 3.36 |
| ... |  |  |  |  |  |  |  |

We then use these summarised data to obtain Mendelian randomisation estimates for each stratum, called localised average causal effect (LACE) estimates, under the assumption that the genetic association with the exposure is constant (“homogeneity assumption”) (8). The LACE estimates are then combined to produce a non-linear function relating the exposure to the outcome using one of two methods: a fractional polynomial method or a piecewise linear method. In the fractional polynomial method, we perform meta-regression of the LACE estimates on the mean values of the exposure within each stratum for a range of parametric models known as fractional polynomials (10). By default, these include linear, quadratic, cubic, logarithmic, reciprocal, and square-root functions. Our package considers fractional polynomials of degree one (one of these functions) or degree two (the sum of two of these functions). This allows a wide range of possible shapes to be fitted to the data. The best-fitting fractional polynomial is chosen using the likelihood function. In the piecewise linear method, we plot a continuous piecewise linear function with slope equal to the LACE estimate in that stratum. In both cases, only the slope of the function is specified, so the function is set to zero at a reference value of the exposure.

The major difference between the individual-level and stratified summarised data versions of the methods is quantification of uncertainty in the model parameters. In the individual-level method, standard errors are estimated in a conventional nonparametric bootstrap by resampling individuals and calculating LACE estimates in the bootstrap samples. In the stratified summarised method, standard errors are estimated in a parametric bootstrap of the LACE estimates by repeatedly drawing from normal distributions with mean at the LACE estimate and standard deviation equal to the standard error of the LACE estimate.

#### Availability

The **SUMnlmr** package can be installed for R version 3.1.0 and higher from https://github.com/amymariemason/SUMnlmr via the command *remotes::install_github(“amymariemason/SUMnlmr”)*. The package dependencies are all CRAN packages: **matrixStats, ggplot2, utils** and **metafor**.

#### Key features

The package facilitates non-linear Mendelian randomisation analyses using stratified summarised data with key features including:

- Creation of stratified summarised data for either a continuous or binary outcome;
- Implementation of the fractional polynomial and piecewise linear methods with bootstrapped confidence intervals;
- Plots of the best-fitting model, with confidence intervals presented either as lines or as a band plot;
- Calculation of test statistics for non-linearity, including 4 tests for the shape of the exposure—outcome relationship: *fp_d1_d2*, a low p-value indicates preference of a degree 2 fractional polynomial compared with a degree 1 fractional polynomial; *fp*, a low p-value indicates preference for a non-linear fractional polynomial model compared with a linear model; *quad*, a low p-value indicates a linear trend in the LACE estimates; and *Q*, a low p-value indicates heterogeneity in the LACE estimates;
- Calculation of test statistics for variability in the genetic associations with the exposure as an assessment of the homogeneity assumption: *Q*, a low p-value indicates heterogeneity in genetic associations; and *trend*, a low p-value indicates a linear trend in the genetic associations.

#### Simulation study

A simulation study was conducted to compare the performance of the fractional polynomial and piecewise linear methods based on individual-level data (implemented using the **nlmr** package, available from https://github.com/jrs95/nlmr) and stratified summarised data (implemented using the **SUMnlmr** package). This simulation study replicates that conducted in the Staley and Burgess paper (4). Results are provided in Supplementary Tables S1-S4. For any given model, coefficient estimates were almost always identical between the two implementations to the first 3 decimal places, and hypothesis tests at a 95% significance level agreed for between 98.4 and 100% of simulated datasets. The only difference between the methods is the individual-level data implementation of the piecewise linear method typically results in a greater extrapolation of the results to a wider range of exposure values, whereas the summarised data implementation results in a more limited range. However, this difference is typically presentational rather than substantive.

### Use

#### LDL-Cholesterol and coronary artery disease

While the causal effect of LDL-cholesterol on coronary artery disease risk is well-established from Mendelian randomisation studies (11) (12) as well as trials of lipid lowering medications (13) (14), the shape of the causal relationship has not been investigated using non-linear Mendelian randomisation methods. We analysed data from UK Biobank on 349,771 individuals of White European ancestry with available data on LDL-cholesterol. The genetic instrument was a weighted score of 87 variants associated with LDL-cholesterol at a genome-wide level of statistical significance in the Global Lipid Genetics Consortium (15). Coronary artery disease was defined using International Classification of Disease (ICD) codes based on routinely collected hospital episode statistics data, death certificates, and self-reported outcomes validated by nurse interview (Supplementary Table S5). Genetic associations with LDL-cholesterol were obtained from linear regression, and with coronary artery disease from logistic regression. Regression models adjusted for age, sex, and the first 10 genetic principal components. Statistical analyses were performed using R version 4.0.3.

Graphs from the fractional polynomial method are displayed in Figure 1 applied to individual-level data (red) and summarised data (blue). Output from the summarised data methods is provided in Supplementary Table S6. Using summarised data, the best-fitting fractional polynomial was a linear function with coefficient 0.32 (95% confidence interval 0.25, 0.38). In the individual data case, the best fit was a quadratic function with coefficient 0.036 (95% confidence interval 0.029, 0.043). However, a linear model fit the data almost as well, and no strong evidence for non-linearity was observed (*p*>0.2 for all tests). The main reason for the difference in results is that the individual-level method fit a model across a wider range of the exposure distribution, and substantial non-linearity was only observed in the extreme tails of the exposure distribution. Our analysis suggests that reductions in LDL-cholesterol will lead to similar proportional reductions in coronary artery disease risk at both low and high levels of LDL-cholesterol.

**Figure 1.**
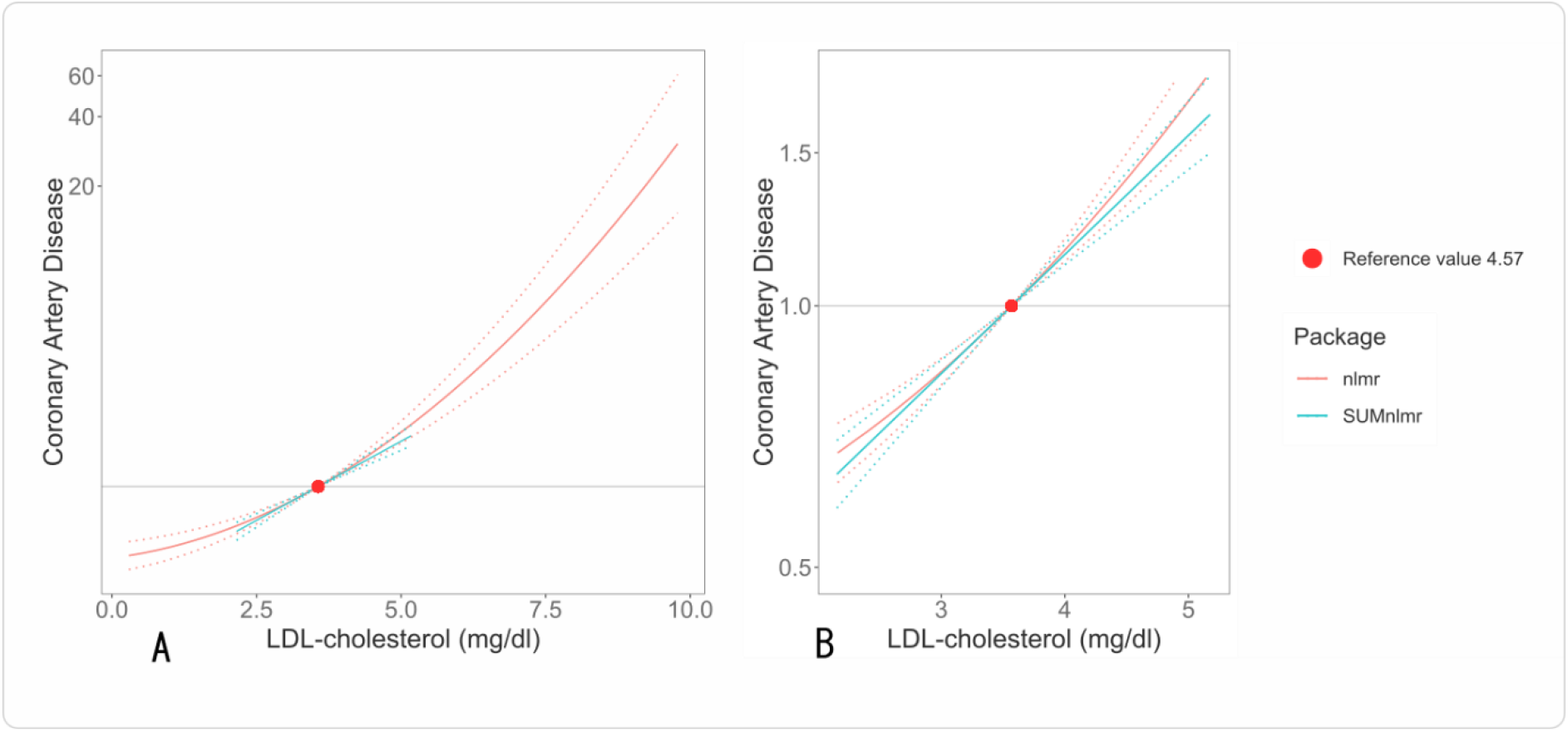
Graph of the best fitting fractional polynomial between LDL-cholesterol and coronary artery disease risk (measured on the odds ratio scale). Panel A shows the models for the range of the exposure distribution considered in the individual-level data version of the method (**nlmr**); Panel B shows the models for the range considered in the summarised data version of the method (**SUMnlmr**);. The range in the individual-level data version of the method is substantially wider due to the presence of individuals with outlying values of LDL-cholesterol. The reference value (4.57 mg/dL) is the mean of LDL-cholesterol in the UK Biobank dataset.

## Discussion

**SUMnlmr** is a software package for R that splits methods for non-linear Mendelian randomisation into two stages: a data manipulation stage that requires access to individual-level data, and a data analysis stage that does not. The two main advantages of this are to facilitate analyses where individual-level data cannot be easily shared, and to separate the customisable part of the analyses from the more technical part of the analysis, allowing the non-linear estimation method to be implemented using the same software code whatever the original form of the individual-level data. Results for a given model choice were generally identical based on individual-level data and stratified summarised data, indicating no loss in power despite the large reduction in the scale of the data. In an applied analysis considering LDL-cholesterol and coronary artery disease, there were differences in model choice between the individual-level and summarised methods, although both indicated no strong evidence that a non-linear model fit the data better than a linear model.

As concerns over the security of individual-level data increase, approaches like this will be required to enable complex analyses without requiring full individual-level data access. While we have framed the two stages of the analysis as being implemented by separate analysts, it could also be that the initial phase of the analysis is performed blindly on a central server (a so-called “walled garden”). The stratified summarised data are reported to the main analyst, who can then perform non-linear analyses on their local computer. This also allows greater accessibility to large data sources, as access to obtain stratified summarised data could be given to a wider set of users. Another context where this could be helpful is facilitating meta-analysis; separate analysts can generate stratified summarised data from their local dataset, and then share these with a central analyst, who can perform complex statistical analyses without ever seeing any individual-level data. Such statistical approaches both accelerate the democratisation of science, by widening access to large datasets, and promote open scientific practice.

## Supporting information

Supplementary materialss

## Data Availability

Individual level data from UK Biobank cannot be shared publicly for ethical/privacy reasons. This data will be shared on reasonable request to the corresponding author, with the permission of UK Biobank.
The stratified summarised data used in the example is in the supplementary material.

https://biobank.ctsu.ox.ac.uk/

## Ethics approval

This research was conducted according to the principles expressed in the Declaration of Helsinki. The UK Biobank cohort has been approved by the North West Multicentre Research Ethics Committee, UK (Ref: 16/NW/0274). Written informed consent has been obtained from all study participants. The current study was approved by the UK Biobank access management board under application 7439. Participants who had withdrawn consent by the time of the analysis were excluded from the dataset.

## Author contributions

Study conception and design: AMM & SB; data creation and analysis: AMM; drafting the manuscript and figures: AMM; reviewing the manuscript: SB & AMM

## Data Availability Statement

Individual level data from UK Biobank cannot be shared publicly for ethical/privacy reasons. The data will be shared on reasonable request to the corresponding author, with the permission of UK Biobank.

The stratified summarised data used in the example is in the online supplementary material.

## Supplementary data

Supplementary data are available at IJE online

