## Supplementary materialss for "Software Application Profile: SUMnlmr, an R package that facilitates flexible and reproducible non-linear Mendelian randomisation analyses"

### Supplementary information for Software Application Profile: SUMnlmr, an R package that facilitates flexible and reproducible non-linear Mendelian randomization analyses

Amy M. Mason, PhD^1,2^, Stephen Burgess, PhD^1,2,3^

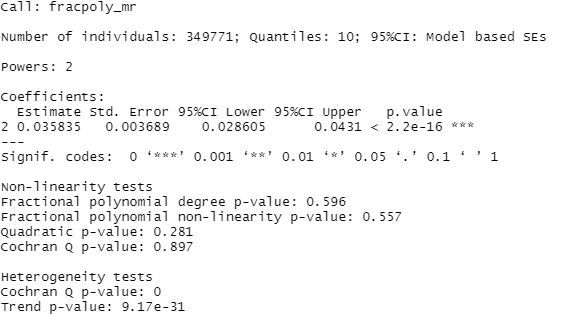

Supp. Figure 1 Output of SUMnlmr for fractional method on the LDL & CAD dataset. Although the results of heterogeneity tests are concerning, as this indicates that the genetic associations with the exposure vary between strata, the extreme low p-values reflect the precision of these estimates. The absolute magnitude of differences between the association estimates was not substantial, particularly when discounting the highest and lowest strata, which are most affected by outliers. As the non-linear method is robust to mild heterogeneity in these associations, we are not overly worried about the validity of results in spite of strong evidence for violation of the homogeneity assumption.

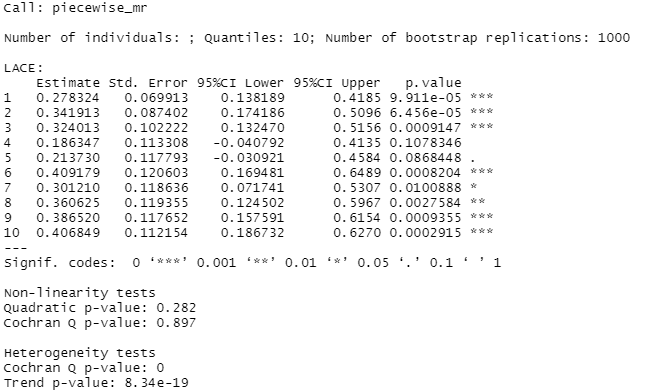

Supp. Figure 2 Output of SUMnlmr for piecewise linear method on the LDL & CAD dataset
